# Outcome Assessment in Neurocritical Care Trials

**DOI:** 10.1101/2023.03.03.23286671

**Authors:** Emily Fitzgerald, Lachlan Donaldson, Oliver Flower, Naomi Hammond, Kwan Yee Leung, Gabrielle McDonald, Kirsten Rowcliff, Ruan Vlok, Anthony Delaney

## Abstract

**Introduction:** The assessment of patient reported outcomes following neurological injury remains a challenging area of neurocritical care research. Mortality amongst the neurocritical patient population remains high with a significant proportion of survivors left suffering functional, cognitive and emotional deficits, often with a reduced health-related quality of life and leaving them dependent on caregivers. Numerous instruments have been developed to assess the level of impairment patients experience following a global neurological injury. Previous systematic reviews have reported significant heterogeneity in outcome assessment in neurocritical car trials, including the outcome measure used, method of ascertainment and the timing of outcome assessment. It has been suggested that this heterogeneity in outcome assessment has complicated the design of neurocritical care clinical trials, the pooling and meta-analysis of trial data and has led to conflicting and controversial trial results. It is unclear what impact the methods of performing outcome assessment has on loss follow up rates and the validity of outcome data in neurocritical care trials.

We aim to systematically review the methods of performing outcome assessment in neurocritical care trials to identify current trends in outcome assessment in this patient population and to examine loss to follow up rates and factors impacting cohort attrition. It is hoped that an understanding of the relationship between methods of outcome assessment and loss to follow up will inform future design of neurocritical care trials.

**Methods and analysis:** This systematic review will include randomized clinical trials and large prospective observational cohort studies where the included population is adults with a diagnosis of traumatic brain injury or a subarachnoid haemorrhage and reporting at least one patient reported outcome measure. Inclusion will not be limited based on intervention nor comparator. We will limit the searches to human studies, with reports published in the English language and published within the last 10 years. We will search the Medline, EMBASE, and The Cochrane Central registry of clinical trial (CENTRAL) for eligible trials. We will manually search the reference list of relevant primary review articles, clinical registries, and abstracts from recent relevant conferences

**Conclusion:** This systematic review and will provide clinicians with an understanding of the relationship between methods of outcome assessment and loss to follow up will inform future design of neurocritical care trials.

## INTRODUCTION

The assessment of patient reported outcomes following neurological injury remains a challenging area of neurocritical care research. Mortality amongst the neurocritical patient population remains high at 5 - 34% (1-3) with a significant proportion of survivors left suffering functional, cognitive and emotional deficits, often with a reduced health-related quality of life and leaving them dependent on caregivers (4, 5). Numerous instruments have been developed to assess the level of impairment patients experience following a global neurological injury (6-8). A systematic review by Tate et al. (2013) identified 728 unique instruments used for outcome assessment in traumatic brain injury (TBI) trials (6) while a systematic review by Andersen et al (2018) reported 285 unique outcome measures, including various functional outcome measures, used in subarachnoid haemorrhage (SAH) trials (8). These reviews reported significant heterogeneity in outcome assessment, including the outcome measure used, method of ascertainment and the timing of outcome assessment (6, 8, 9). It has been suggested that this heterogeneity of outcome assessment has complicated the design of neurocritical care clinical trials, the pooling and meta-analysis of trial data and has led to conflicting and controversial trial results (4, 9-11).

It is unclear what impact the methods of performing outcome assessment has on loss follow up rates and the validity of outcome data in neurocritical care trials. Neurocritical care trials often report high loss to follow up rates of 15-29% (2, 12-14). Although inevitable in long term follow up studies, loss to follow up can bias trial results (15, 16). It has been suggested that a loss to follow-up of <5% leads to little bias while >20% poses a threat to the validity of trial results (15, 17).

### Objectives

To identify methods of outcome assessment and examine loss to follow up in trials published between 2010 and 2020 enrolling patients admitted to intensive care with aneurysmal SAH or TBI.

## METHODS

We will conduct a systematic review of randomised clinical trials and prospective cohort studies in accordance with the recommendations of the Cochrane Handbook of Systematic Reviews of Interventions. This systematic review has been registered on the International Prospective Register of Systematic Reviews

### ELIGIBILITY CRITERIA

#### Study Types

This systematic review will include all randomized clinical trials and prospective observational cohort studies that included a minimum of 100 participants.

#### Population

We will include trials where the included population is adults with a diagnosis of TBI or aSAH

#### Intervention/comparator

Inclusion will not be limited based on intervention nor comparator.

#### Outcomes

We will include studies reporting at least one patient reported outcome measure.

#### Exclusion criteria

We will limit the searches to human studies, with reports published in the English language and published within the last 10 years

### Information Sources

We will search the Medline, EMBASE, and The Cochrane Central registry of clinical trial (CENTRAL) for eligible trials, if any. We will manually search the reference list of relevant primary review articles, clinical registries, and abstracts from recent relevant conferences and contact experts in the field.

### Search Strategy

We will search Medline and EMBASE (using the OVID interface) and CENTRAL. We will conduct MeSH and keyword searches for aneurysmal subarachnoid haemorrhage or traumatic brain injury combined with MeSH and keyword searches for intensive care or critical care, with filters for randomized clinical trials and observational studies.

## DATA MANAGEMENT

References of studies yielded by the search will be uploaded into COVIDENCE. Data will be extracted into a purpose built excel spreadsheet and analysed in STATA.

### Study Selection process

The review authors will develop screening forms based on the eligibility criteria. To ensure consistency between reviewers, a calibration exercise will be undertaken to pilot and refine the screening forms prior to commencing the formal screening process.

The review authors (EF, QF, RV, GM and KR) will independently and in duplicate screen the titles and abstracts yielded by the search. Full-text reports will be obtained for all titles and abstracts that appear to meet the eligibility criteria or where there is any uncertainty. The review authors will independently screen the full-text articles and decide whether they meet the eligibility criteria. We will seek additional information from study authors where necessary to resolve questions about eligibility. Reviewers will resolve disagreements by discussion, and an arbitrator will adjudicate unresolved disagreements.

### Data Collection Process

The review authors will develop data collection forms and a detailed instruction manual to extract data from included studies. To ensure consistency between reviewers, a calibration exercise will be undertaken to pilot and refine the data collection form prior to commencing the formal data collection process. The review authors will extract data independently and in duplicate from each included study. We will seek additional information from study authors where necessary to resolve questions. Reviewers will resolve disagreements by discussion, and an arbitrator will adjudicate unresolved disagreements.

### Data Items

We will extract data regarding study characteristics (including first author, year of publication, study type, number of participants, location of site, number of sites) as well as details of patient outcome assessment (primary outcome measure, patient reported outcome measure used, method of ascertainment, blinding of outcome assessors, training of outcome assessors, timing of follow up, duration of follow up, loss to follow up)

### Risk of bias individual studies

The review authors will independently and in duplicate make a judgement as to the possible risk of bias of each included study based on various domains. If there is insufficient detail reported in the study, we will judge the risk of bias as ‘unclear’. Reviewers will resolve disagreements by discussion, and an arbitrator will adjudicate unresolved disagreements.

The review authors will assess the quality of included RCT using the Cochrane Collaboration tool to assess risk of bias for randomised trials. The following domains will be addressed in assessing the risk of bias:

- Selection bias (including method of randomization and allocation concealment)
- Performance bias
- Detection Bias
- Attrition Bias
- Selective reporting bias
- Other bias

The review authors will assess the quality of included prospective cohort studies using the Newcastle-Ottawa for assessing the risk of bias. The following domains will be addressed in assessing the risk of bias:

- Bias due to confounding
- Bias in selection of participants into the study
- Bias in classification of intervention
- Bias due to deviation from intended interventions
- Bias in measurement of outcomes
- Bias in selection of reported result

### OUTCOMES

#### Primary

To describe patient reported outcome assessment in neurocritical care trials, specifically;

- Patient reported outcome measure/s used
- Methods of assessing patient reported outcome

#### Secondary

To report other key aspects of outcome assessment in neurocritical care trials, specifically;

- Primary outcome measure of the trial
- Timing and duration of outcome assessment
- Loss to follow up rates

## Data Availability

All data produced in the present study are available upon reasonable request to the authors

## DATA SYNTHESIS

We will present simple statistics regarding the range and frequency of use of patient reported outcome assessment instruments, method of assessment, timing of outcome measurement.

## ETHICS AND DISSEMINATION

This review does not require ethical approval as this is a systematic review of published studies. We plan to present the results of the systematic review at national and international scientific meetings and will prepare a manuscript for submission to a peer reviewed journal.

## DISCUSSION AND LIMITATIONS

This systematic review will provide a comprehensive review of patient reported outcome assessment in neurocritical care trials. We acknowledge that there will be limitations to the proposed systematic review, including that eligible studies are anticipated to be heterogeneous in nature due to variations in the included trials, such as the trial design, trial population and the intervention or comparator used. In addition, the strength of the systematic review may be limited by the quality of reporting of the outcome assessment in included studies.

